# Socio-economic Transition in an Indian Rural Setting and Its Impact on Health: Pune Maternal Nutrition Study

**DOI:** 10.1101/2021.07.06.21260054

**Authors:** S R. Shukla, K Kumaran, A Radkar, A J. Bhalerao, V A. Solat, S D. Chougule, T M. Deokar, P C. Yajnik, C HD. Fall, C S. Yajnik

## Abstract

**Background:** There is scant scientific information on socioeconomic transition and its effects on health parameters in India. The prospective data collection in the Pune Maternal Nutrition Study (PMNS) birth cohort provided an opportunity to examine the changes in socio-economic status (SES) and 1) its comparison against state and district indicators, 2) association with body size and cardio-metabolic risk factors.

**Method:** We followed up ∼700 families for biomedical information in the PMNS. SES data was collected in the index pregnancy (1993-96) and serially every six years (∼2001, ∼2007, ∼2013). Comparisons with rural Maharashtra and Pune were made using data from the National Family Health Survey 1999 and 2006 (NFHS II and III).

**Results:** There was an increase in the proportion of those in the high Standard of Living Index (SLI, NFHS) from 64% to 90% between 2001 and 2013. The increase in SES in our study area was faster and greater compared to the state and district. Higher SLI was associated with higher weight, height, body fat percent and total cholesterol concentrations at each followup. However, there were no associations between transition and health parameters.

**Conclusion:** Our data suggest a rapid socio-economic transition in the study area which is faster than the state and district.

## Introduction

India is undergoing rapid socio-economic changes accompanied by an epidemiological transition. While communicable diseases exist, rates of diabetes and cardiovascular disease are rising rapidly^1,2,3,4^. This has been partly attributed to greater urbanisation, adoption of energy dense diets and, lower levels of physical activity^5,6^. However, there is sparse data to quantify the socio-economic transition that has occurred recently.

Socio-economic transition can be measured by examining changes in socio-economic status (SES). SES indices change with time and each index uses parameters that are considered appropriate at that point. In community, based studies in India, the Parikh-Trivedi scale ^7^, Kuppuswamy scale,^8^ or Standard of Living Index^9^ have all been used over the years. Although some of the parameters used may be different, all these scales measure related aspects.

We had a unique opportunity to study socio-economic change and its impact on health parameters in a rural Indian population where serial data on socio-economic indicators and health were collected at various time points between 1993-95 and 2013-14 (the Pune Maternal Nutrition Study {PMNS}). We aimed to assess the socio-economic transition in this cohort and examine its effect on health outcomes by:

- Analysing the changes in SES in the study cohort to assess any transition
- Comparing this change with that occurring in rural Pune district and Maharashtra state to assess relative pace of transition
- Examining the associations between change in SES and health status using anthropometry and biochemical indicators.

### Research Design and Methods

The methodology of the PMNS has been published earlier ^9,10^. In brief, it was set up in 1993 in 6 villages near Pune to study the influence of maternal nutrition on fetal growth and later cardiovascular risk of the offspring. Married non-pregnant women were enrolled and monitored for pregnancy; 797 women with a singleton pregnancy entered the study. Their offspring were subsequently followed up at birth and 6, 12 and 18 years for detailed anthropometric and cardio-metabolic measurements along with dietary intake.

Ethical approval for the study was obtained from the Ethics Committee of KEM Hospital Research Centre and informed consent was obtained from all participants.

### Measurements on the offspring

The offspring underwent anthropometry at birth and detailed cardio-metabolic assessments at 6, 12 and 18 years of age. A 75g oral glucose tolerance test (OGTT; WHO criteria) was carried out at 6 and 18 years, while a single fasting blood sample was collected at 12 years. Anthropometric measurements included height, weight, and skinfolds. Body composition was measured using dual X-ray absorptiometry (DXA). Blood pressure was recorded using standardised methods. Blood was analysed for plasma glucose, total cholesterol, HDL-cholesterol, and triglycerides using standard enzymatic kits.

### Socio-economic status assessment

At baseline and 6 years, SES was assessed using the Parikh and Trivedi weighted score which considers education, occupation, family type, type of house, land ownership, agricultural equipment and material possessions. At 6, 12 and 18years we used the Standard of Living Index (SLI) to calculate household SES. The SLI was calculated using the method followed in the National Family Health Survey (NFHS) rounds 2 (1998-9) and 3 (2005-6)^11, 12^. The index is composed of information on type of house, toilet facility, source of lighting, fuel for cooking, source of drinking water, and ownership of house, agricultural land, irrigated land, livestock, and material goods. Weights are assigned for individual components and a composite score is computed. Data were available on 803 households at baseline (1994-95), 677 at 6 years (2001-02), 596 at 12 years (2007-08), and 659 (2013-14) at 18 years.

To study relative pace of transition, we used comparative figures for SES at state (NFHS II 1998 and NFHS III 2006) and district and village level (Census 1991 and 2001) using survey data^13,14,15,16,17^.

### Statistical analysis

We assessed transition by describing and comparing the change in SLI parameters from baseline to 6, 12, and 18 years and between each of these periods. We also compared the transition in our cohort with the district (Pune) and state (Maharashtra).

We assessed the relationship between SES and health parameters using correlation coefficients. We calculated the changes in SLI scores between 6 and 18 years, and 12 and 18 years and correlated it with anthropometric and biochemical parameters. We further divided the cohort into groups based on the amount of change in SES scores (no change, below and above mean) and compared health parameters across these groups. We also assessed the relationship between health parameters at 12 years and change in SLI scores between 6 and 12 years in the mothers (mean age 33y in mothers).

## Results

### SES transition in the study area

The study villages have undergone considerable changes in the infrastructure during last 20 years. ((Supplementary Table 1) ^14-15^). Originally described as drought-prone, villages have seen an increase in irrigated land with two dams (1994 and 2001) enabling irrigation throughout the year. Transport facilities have improved increasing access for work and education outside the villages. Local primary care centres have improved access to health care. New higher education institutes have started locally increasing access to education. All the villages now have banking facilities. The government has supported the development of two industrial belts in the area resulting in increased employment opportunities.

**Table 1:**
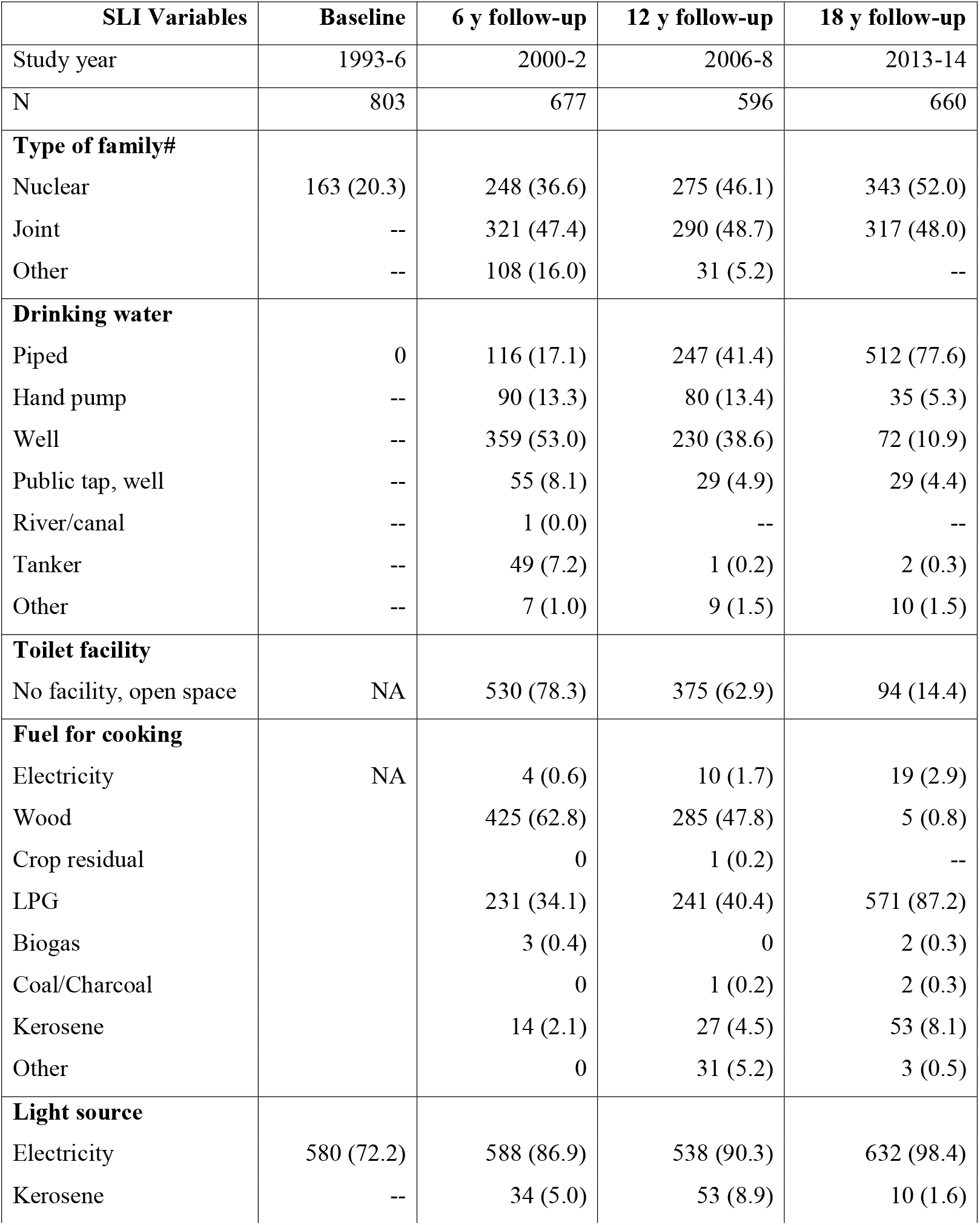
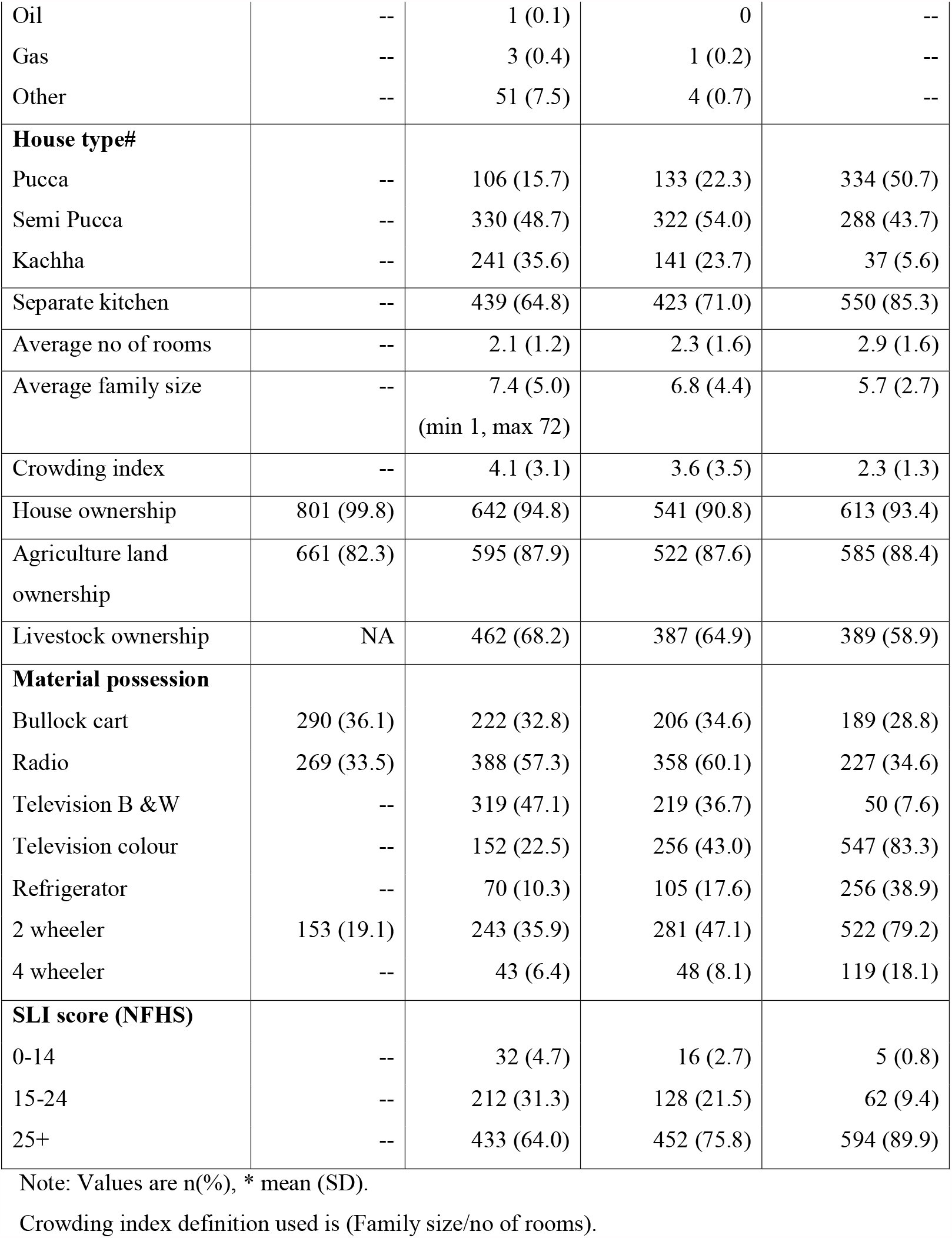
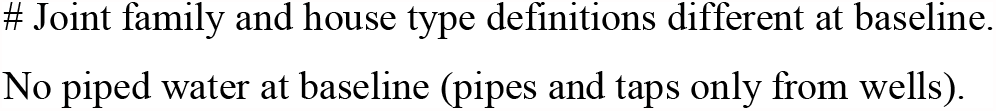
Standard of living index (SLI) at the 6, 12 and 18 years follow-up of the study cohort.

Table 1 shows that between baseline and 6years, there was an increase in nuclear families (from 20% to 36%). The proportion of those who were cultivators decreased (from 68% to 53%) with an increase in those engaged in service (from 9 to 18%). There was also an overall increase in material possessions, reflecting in higher SLI scores. Using the NFHS classification of SLI categories, the proportion of those in the high SLI category increased from 64% at 6 years to 77% at 12 and 90% at 18 years. The proportion of those in the low SLI category is ∼1%.

The proportion of *pucca* and *semi-pucca* houses (built using robust/permanent material) increased while that of *kachha* houses (built using flimsy non-permanent material) decreased; this change was marked between 12 and 18 years. A rise in the number of rooms in households accompanied by a decrease in the number of people is reflected in a lower crowding index. Supply of safe piped water increased from nil at the baseline to 17%, 42% and 78% serially till 18 years. The proportion of households using well water declined from ∼100 % at baseline to 53% at 6 and 11% at 18 years. Toilet facilities improved overall; the change was greatest between 12 and 18 years with access to toilets increasing by ∼48%. The use of LPG as the main cooking fuel and availability of electricity increased overall; currently 90% use smokeless fuel for cooking and 98% of households have electricity. The change was substantial between 12 and 18 years.

There was little change in the proportion of households owning land but the ownership of household consumer goods including refrigerators, colour TVs, two-wheelers and cars progressively increased, particularly between 12 and 18 years, so that 79% of households own a two-wheeler and 18% own four-wheelers.

The percentage of women marrying before 18 years was 46.7% at baseline in 1993-94, but only ∼13% in their daughters. This has reflected in higher educational attainments. Ten percent of the mothers were illiterate, and only 10% had reached 10^th^ standard in the school. In contrast, majority (81%) of daughters have studied beyond 10 years, and 19% are in graduate or professional courses.

### Comparison of SES transition between the study area, and the state and district

The distribution of SLI scores in the study area and rural Maharashtra (Figure 1) suggests that there has been an overall shift of people into the highest SLI category; however, the proportion of those in the highest SLI category is substantially higher in the study area. The change in mean scores was also greater in the study area indicating a higher pace of transition.

**Fig 1:**
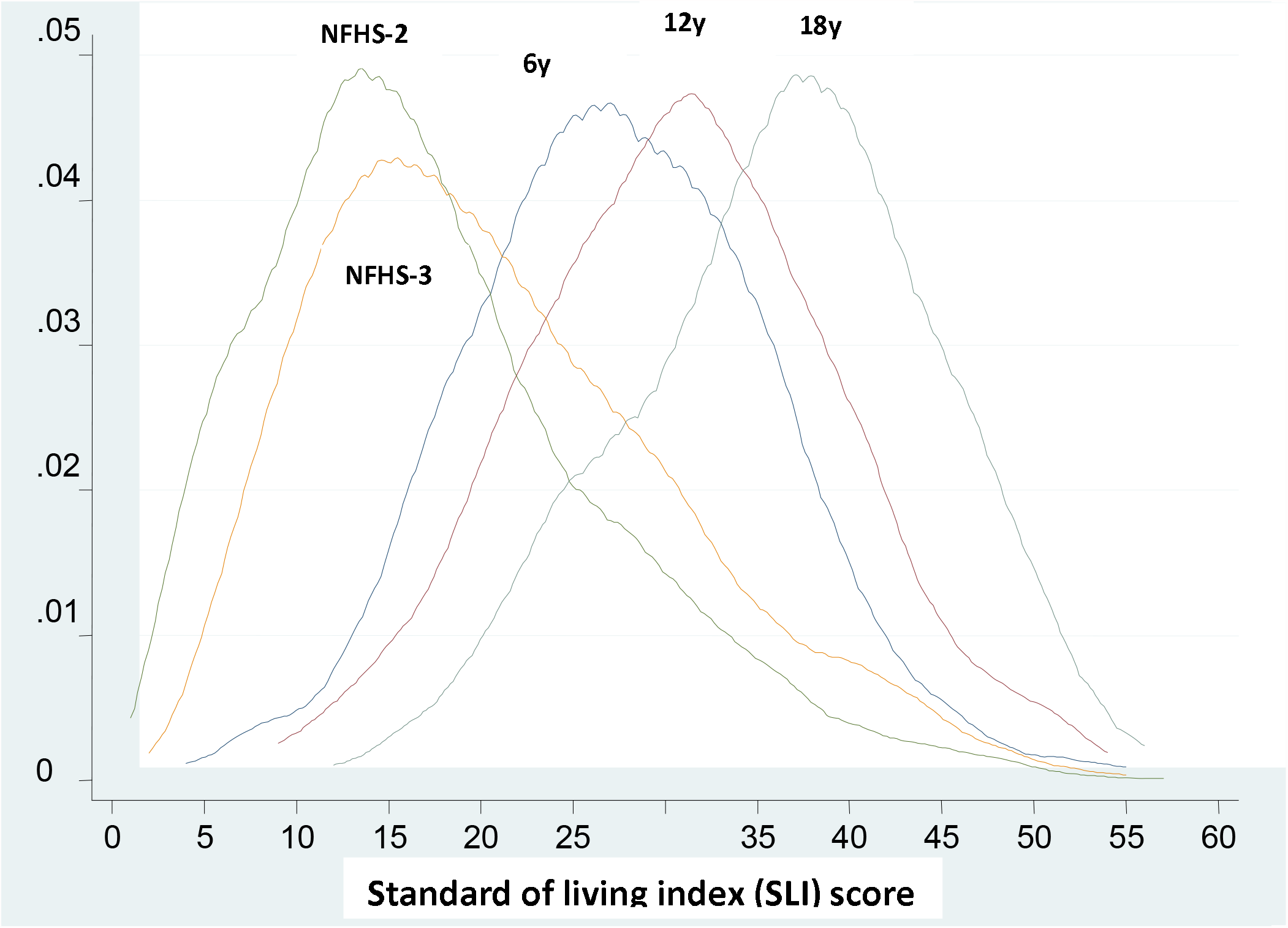
Distribution of SLI scores in the study area at the 6(2000-2), 12 (2006-8) and 18 years (2013-14) follow-up compared with rural Maharashtra at the NFHSII (1998-9) and NFHS III (2005-6) surveys.

Table 2 compares data on different development parameters and SES indicators for the study area with the rural areas for the state (Maharashtra), and the district (rural Pune), for broadly comparable time periods.

**Table 2:** Percent distribution of households by socioeconomic characteristics of the study cohort (6, 12 and 18 years follow-up) compared with rural Maharashtra and Pune.

|  | PMNS cohort |  |  |  |  |  | Rural Maharashtra |  |  | Rural Pune |  |  |
| --- | --- | --- | --- | --- | --- | --- | --- | --- | --- | --- | --- | --- |
| Variables | 6y | 12y | 18y | Δ for %<br>change<br>(6-12y) | Δ for %<br>change<br>(12-18y) | Δ for %<br>change<br>(6-18y) | NFHS II | NFHS III | Δ for %<br>change | Census | Census | Δ for %<br>change |
| **Year of study** | 2000-02 | 2006-08 | 2013-14 |  |  |  | 1998-9 | 2006-7 |  | 2001 | 2011 |  |
| Number of household | 570 | 570 | 659 |  |  |  | 3298 | 2466 |  |  |  |  |
| **Type of family** |  |  |  |  |  |  |  |  |  |  |  |  |
| Nuclear | 36.1 | 45.6 | 52.0 | 26.3 | 12.3 | 44.0 | 54.3 | 56.6 | 4.2 |  |  |  |
| **House** |  |  |  |  |  |  |  |  |  |  |  |  |
| House ownership | 94.9 | 91.4 | 93.4 | (-)3.7 | 2.2 | (-)1.6 | 91.9 | 92.4 | 0.5 |  |  |  |
| **House type** |  |  |  |  |  |  |  |  |  |  |  |  |
| Pucca | 16.1 | 22.3 | 50.7 | 38.5 | 127.3 | 214.9 | 9.5 | 30.9 | 125.3 |  |  |  |
| Semi Pucca | 47.5 | 54.0 | 35.4 | 13.6 | (-)34.4 | (-)25.5 | 21.4 | 63.9 | 198.6 |  |  |  |
| Kachha | 36.3 | 23.7 | 4.5 | (-)34.7 | (-)81.0 | (-)87.6 | 68.9 | 4.9 | (-)92.8 |  |  |  |
| **Drinking water** |  |  |  |  |  |  |  |  |  |  |  |  |
| Piped | 17.0 | 41.9 | 77.6 | 146.4 | 85.2 | 356.5 | 22.5 | 37.0 | 64.4 | 47.5 | 51.4 | 9.4 |
| Well | 53.5 | 38.2 | 10.9 | (-)28.6 | (-)71.5 | (-)79.6 | 26.1 | 16.4 | (-)37.2 | 33.6 | 24.2 | (-)28.0 |
| **Toilet facility** |  |  |  |  |  |  |  |  |  |  |  |  |
| Open space | 78.1 | 62.8 | 14.4 | (-)19.6 | (-)77.1 | (-)81.6 | 85.0 | 79.3 | (-)6.5 | 81.5 | 38.2 | (-)53.1 |
| **Cooking fuel type** |  |  |  |  |  |  |  |  |  |  |  |  |
| LPG/natural gas | 33.7 | 40.7 | 87.2 | 20.8 | 114.2 | 158.7 | 9.9 | 16.1 | 76.9 | 17.0 | 49.0 | 188.2 |
| **Light source** |  |  |  |  |  |  |  |  |  |  |  |  |
| Electricity | 87.2 | 90.5 | 98.4 | 3.8 | 8.7 | 12.8 | 71.4 | 70.5 | (-)1.3 | 75.4 | 83.3 | 10.5 |
| **Agricultural land** |  |  |  |  |  |  |  |  |  |  |  |  |
| Land ownership | 88.9 | 88.6 | 88.4 | (-)0.3 | (-)0.2 | (-)0.6 | 66.9 | 57.9 | (-)13.5 |  |  |  |
| **Material possession** |  |  |  |  |  |  |  |  |  |  |  |  |
| Bicycle | 77.0 | 86.1 | 80.9 | 11.8 | (-)6.0 | 5.1 | 35.3 | 37.8 | 7.4 | 42.6 | 39.7 | (-)6.8 |
| Refrigerator | 10.3 | 17.6 | 38.9 | 70.8 | 121.0 | 277.7 | 4.0 | 6.1 | 52.5 |  |  |  |
| 2 wheeler | 36.0 | 47.7 | 79.2 | 32.5 | 66.0 | 120.0 | 8.4 | 13.1 | 55.9 | 16.3 | 38.7 | 137.4 |
| 4 wheeler | 5.8 | 8.1 | 18.1 | 39.6 | 123.4 | 212.1 | 0.8 | 1.1 | 37.5 | 3.3 | 6.0 | 81.8 |
| **SLI** |  |  |  |  |  |  |  |  |  |  |  |  |
| Low | 4.9 | 2.3 | 0.8 | (-)53.1 | (-)65.2 | (-)83.7 | 47.6 | 34.2 | (-)28.1 |  |  |  |
| Medium | 30.7 | 20.9 | 9.4 | (-)31.9 | (-)55.0 | (-)69.4 | 40.8 | 35.4 | (-)13.2 |  |  |  |
| High | 64.4 | 76.8 | 89.9 | 19.3 | 17.1 | 39.6 | 9.8 | 30.4 | 210.2 |  |  |  |

**Values are percent. Total will not add upto 100 as some minor categories are not shown. A blank indicates information not available.**
**Collected SLI data used for study area; NFHS data used for rural Maharashtra; Census data used for rural Pune.**

The proportion of nuclear families in the study area was lower in comparison to rural Maharashtra although the percentage change was higher; there are no comparable data available for rural Pune. While *kachha* houses have decreased with a corresponding increase in *pucca and semi-pucca* houses, the change is greater in rural Maharashtra than in the study area

At baseline, there was no access to piped water in the study area, now it exceeds that in the rest of rural Maharashtra. The magnitude of change was also larger in the study area. At baseline, the study area had better access to toilet facilities compared to rural Maharashtra. Toilet facilities have increased considerably in the study area with over 85% having access to closed toilet facilities; this figure and the magnitude of change is higher than that for rural Maharashtra and Pune. The availability of LPG as cooking fuel and electricity as lighting source were higher in the study area compared to rural Maharashtra and Pune at baseline and continue to be higher (Supplementary figure 1). The study area had higher level of material possessions at baseline and a steeper increase compared to rural Maharashtra and Pune.

### Relationship with health parameters

At 6 years, children with higher SLI scores were taller, heavier and had greater weight gain from birth. They also had higher head and mid-arm circumferences, and waist-hip ratio after adjusting for age at the time of measurement, gender and parity (p<0.05 for all). Similar findings were observed 12 and 18 years. At 6 years, children with higher SLI scores had higher total cholesterol concentrations, and lower inflammatory markers (total white cell count). At 12 years, those with higher SLI scores had higher total cholesterol levels while at 18 years, they had higher haemoglobin and triglycerides. Blood pressure, fasting, and 2h glucose and insulin concentrations were unrelated to SLI scores at any age. Dietary intake was available at 12 and 18 years; children with higher SLI scores had higher total caloric (2089 vs 1958 k cals), fat (41 vs 37) and protein intake (58 vs 52 g) at 12 years compared to those children from lower SES (p<0.01 for all). Results are similar at 18y.

We also examined the effects of socio-economic transition on anthropometry and biochemical parameters on the children by considering the change between 6 and 18 (Table 3), 6 and 12 (Supplementary Table 2), and 12 and 18 years (Supplementary Table 3). There were no clinically or statistically significant relationships between transition and health parameters.

**Table 3:**
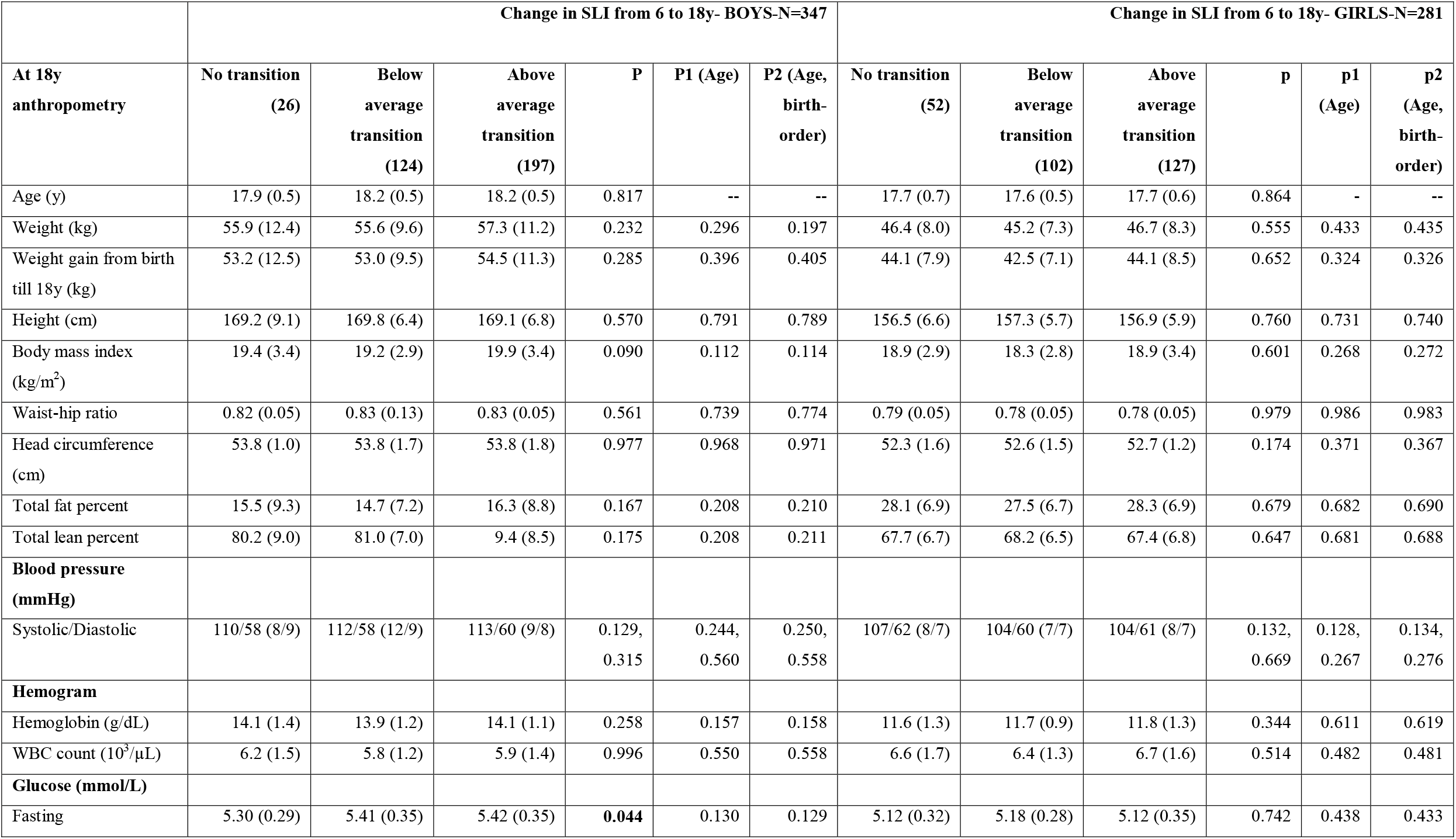
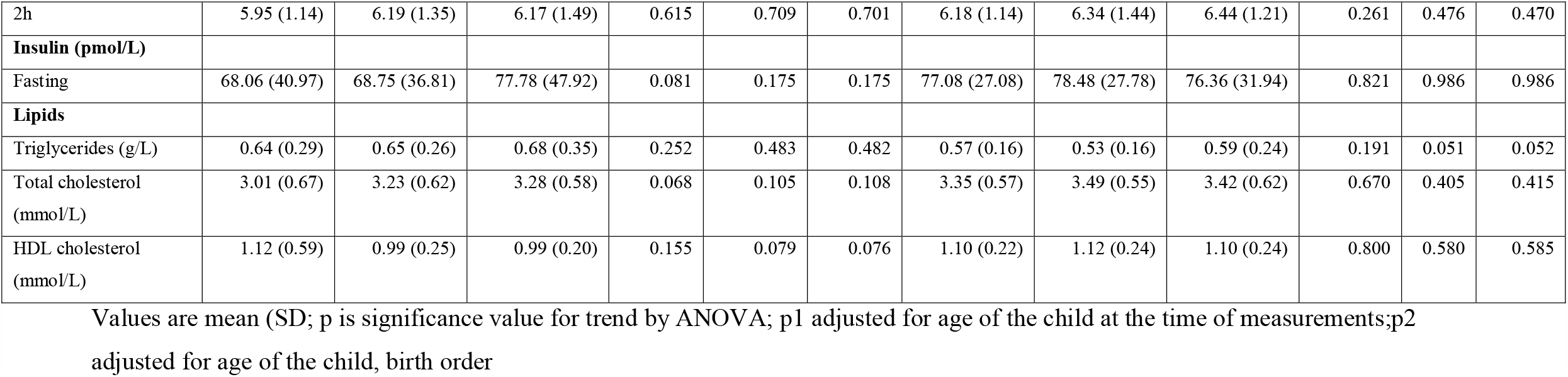
Child anthropometry, biochemistry, and nutritional intake at 18 years follow-up by change (6 to 18 years) in Standard of Living Index, according to gender.

|  | Change in SLI from 6 to 18y- BOYS-N=347 |  |  |  |  |  | Change in SLI from 6 to 18y- GIRLS-N=281 |  |  |  |  |  |
| --- | --- | --- | --- | --- | --- | --- | --- | --- | --- | --- | --- | --- |
| At 18y anthropometry | No transition<br>(26) | Below average transition<br>(124) | Above average transition<br>(197) | P | P1 (Age) | P2 (Age, birth-order) | No transition<br>(52) | Below average transition<br>(102) | Above average transition<br>(127) | p | p1 (Age) | p2 (Age, birth-order) |
| Age (y) | 17.9 (0.5) | 18.2 (0.5) | 18.2 (0.5) | 0.817 | -- | -- | 17.7 (0.7) | 17.6 (0.5) | 17.7 (0.6) | 0.864 | - | -- |
| Weight (kg) | 55.9 (12.4) | 55.6 (9.6) | 57.3 (11.2) | 0.232 | 0.296 | 0.197 | 46.4 (8.0) | 45.2 (7.3) | 46.7 (8.3) | 0.555 | 0.433 | 0.435 |
| Weight gain from birth till 18y (kg) | 53.2 (12.5) | 53.0 (9.5) | 54.5 (11.3) | 0.285 | 0.396 | 0.405 | 44.1 (7.9) | 42.5 (7.1) | 44.1 (8.5) | 0.652 | 0.324 | 0.326 |
| Height (cm) | 169.2 (9.1) | 169.8 (6.4) | 169.1 (6.8) | 0.570 | 0.791 | 0.789 | 156.5 (6.6) | 157.3 (5.7) | 156.9 (5.9) | 0.760 | 0.731 | 0.740 |
| Body mass index (kg/m<sup>2</sup>) | 19.4 (3.4) | 19.2 (2.9) | 19.9 (3.4) | 0.090 | 0.112 | 0.114 | 18.9 (2.9) | 18.3 (2.8) | 18.9 (3.4) | 0.601 | 0.268 | 0.272 |
| Waist-hip ratio | 0.82 (0.05) | 0.83 (0.13) | 0.83 (0.05) | 0.561 | 0.739 | 0.774 | 0.79 (0.05) | 0.78 (0.05) | 0.78 (0.05) | 0.979 | 0.986 | 0.983 |
| Head circumference (cm) | 53.8 (1.0) | 53.8 (1.7) | 53.8 (1.8) | 0.977 | 0.968 | 0.971 | 52.3 (1.6) | 52.6 (1.5) | 52.7 (1.2) | 0.174 | 0.371 | 0.367 |
| Total fat percent | 15.5 (9.3) | 14.7 (7.2) | 16.3 (8.8) | 0.167 | 0.208 | 0.210 | 28.1 (6.9) | 27.5 (6.7) | 28.3 (6.9) | 0.679 | 0.682 | 0.690 |
| Total lean percent | 80.2 (9.0) | 81.0 (7.0) | 9.4 (8.5) | 0.175 | 0.208 | 0.211 | 67.7 (6.7) | 68.2 (6.5) | 67.4 (6.8) | 0.647 | 0.681 | 0.688 |
| **Blood pressure (mmHg)** |  |  |  |  |  |  |  |  |  |  |  |  |
| Systolic/Diastolic | 110/58 (8/9) | 112/58 (12/9) | 113/60 (9/8) | 0.129, 0.315 | 0.244, 0.560 | 0.250, 0.558 | 107/62 (8/7) | 104/60 (7/7) | 104/61 (8/7) | 0.132, 0.669 | 0.128, 0.267 | 0.134, 0.276 |
| **Hemogram** |  |  |  |  |  |  |  |  |  |  |  |  |
| Hemoglobin (g/dL) | 14.1 (1.4) | 13.9 (1.2) | 14.1 (1.1) | 0.258 | 0.157 | 0.158 | 11.6 (1.3) | 11.7 (0.9) | 11.8 (1.3) | 0.344 | 0.611 | 0.619 |
| WBC count (10<sup>3</sup>/μL) | 6.2 (1.5) | 5.8 (1.2) | 5.9 (1.4) | 0.996 | 0.550 | 0.558 | 6.6 (1.7) | 6.4 (1.3) | 6.7 (1.6) | 0.514 | 0.482 | 0.481 |
| **Glucose (mmol/L)** |  |  |  |  |  |  |  |  |  |  |  |  |
| Fasting | 5.30 (0.29) | 5.41 (0.35) | 5.42 (0.35) | **0.044** | 0.130 | 0.129 | 5.12 (0.32) | 5.18 (0.28) | 5.12 (0.35) | 0.742 | 0.438 | 0.433 |
| 2h | 5.95 (1.14) | 6.19 (1.35) | 6.17 (1.49) | 0.615 | 0.709 | 0.701 | 6.18 (1.14) | 6.34 (1.44) | 6.44 (1.21) | 0.261 | 0.476 | 0.470 |
| **Insulin (pmol/L)** |  |  |  |  |  |  |  |  |  |  |  |  |
| Fasting | 68.06 (40.97) | 68.75 (36.81) | 77.78 (47.92) | 0.081 | 0.175 | 0.175 | 77.08 (27.08) | 78.48 (27.78) | 76.36 (31.94) | 0.821 | 0.986 | 0.986 |
| **Lipids** |  |  |  |  |  |  |  |  |  |  |  |  |
| Triglycerides (g/L) | 0.64 (0.29) | 0.65 (0.26) | 0.68 (0.35) | 0.252 | 0.483 | 0.482 | 0.57 (0.16) | 0.53 (0.16) | 0.59 (0.24) | 0.191 | 0.051 | 0.052 |
| Total cholesterol (mmol/L) | 3.01 (0.67) | 3.23 (0.62) | 3.28 (0.58) | 0.068 | 0.105 | 0.108 | 3.35 (0.57) | 3.49 (0.55) | 3.42 (0.62) | 0.670 | 0.405 | 0.415 |
| HDL cholesterol (mmol/L) | 1.12 (0.59) | 0.99 (0.25) | 0.99 (0.20) | 0.155 | 0.079 | 0.076 | 1.10 (0.22) | 1.12 (0.24) | 1.10 (0.24) | 0.800 | 0.580 | 0.585 |

Values are mean (SD; p is significance value for trend by ANOVA; p1 adjusted for age of the child at the time of measurements;p2 adjusted for age of the child, birth order

Mothers with higher SLI scores had greater weight, BMI, total cholesterol and triglyceride concentrations. Mothers who underwent a greater degree of transition (as measured by change in SLI scores) between their child’s 6 and 12 year follow-ups had higher weight, BMI and total cholesterol levels after adjusting for age and initial SLI score (Supplementary Table 4).

## Discussion

This paper describes the transition in SES in a cohort of people from six villages in Pune district, in the state of Maharashtra, India. The cohort was setup in 1993 and the latest follow-up was in 2013-14. During this period, the proportion of households in the high SLI category has progressively increased. The proportion of nuclear families has increased with a reduction in crowding. There has been a substantial increase in the provision of safe drinking water, access to toilets, the use of smokeless fuel and material possessions. Educational attainment of young women showed a marked increase compared with their mothers. These findings indicate socio-economic transition, which has been greatest in the last 6 years.

To compare the transition in our study area with the district and the state, we mainly compared four parameters of development: drinking water, toilet facilities, electricity, and cooking fuel. Comparison with the district (rural Pune) is more relevant whereas comparison with the state (Maharashtra) has inherent constraints. Pune is a developed district of Maharashtra, while the state consists of the developed western region along with the less developed districts from eastern and central areas. Overall the study area is better off in all the four parameters of development under consideration, compared to the state and district. The magnitude of change has also been greater indicating rapidity of transition. This may suggest that the study area has better infrastructure development. Ownership of material possessions is also higher in the study area indicating increased purchasing power. Land ownership remains unchanged in the study area indicating that this group has not experienced financial situations requiring the sale of their land.

Factors that may have contributed to this phenomenon include the availability of better transport and communication links, increased irrigation facilities, and the development of an industrial belt in the vicinity. Improved road conditions, increased public transport facilities, and possession of automobiles make it possible for people to travel to work to the industrial area, or to urban Pune. The industrial belt has increased employment opportunities and many women work outside the home. Availability of better irrigation facilities has allowed farming families to grow cash crops instead of remaining subsistence farmers. Initiatives to improve agricultural technology and farming methods have increased yields. Agricultural produce is also being sent to distant markets with the availability of better transport links. An increasing proportion of younger people are opting for higher education which will lead to further benefits in the longer term.

Our data shows that children with higher SES scores are heavier and taller throughout childhood and into young adulthood. This is reflected in a progressive reduction in the prevalence of stunting and anaemia but higher levels of circulating lipids. However, we were unable to find a statistical association between health parameters and the degree of transition in these children. On the other hand, mothers who had experienced the greatest change in SES had higher weight, BMI, and cholesterol levels. It is possible that for any changes to become apparent, it takes a relatively longer time in children and may become more obvious with increasing age. Homeostatic mechanisms in children are thought to be more effective than in adults and may therefore be a factor for the lack of association between change in SES and health parameters. A recent study in India showed that most cardio-metabolic risk factors evolve gradually when people are exposed to lifestyle changes^18^.

Our study provided a unique opportunity to assess and document the SES transition and its effect on health parameters in a rural cohort with longitudinal data collected over 20 years and across two generations. Data on SES and health were systematically collected by a small research team using standardized methods. This allows us to be confident in our assessment of transition and its effect on health. We acknowledge that we were unable to use the SLI to assess baseline data because this tool became available only later.

Overall, we have highlighted a substantial socioeconomic transition in the study area over the past 18 years, with a sizeable improvement over the last 6 years. We speculate these factors will reflect in changes to health parameters in the longer term.

## Data Availability

The datasets used for this study may be obtained from the corresponding author on reasonable request. The data supply is governed by rules of the Government of India (HMSC).

## List of abbreviations

PMNS: Pune Maternal Nutrition Study
BMI: Body Mass Index
LPG: Liquid Petroleum Gas
NFHS: National Family Health Survey
OGTT: Oral Glucose Tolerance Test
SES: Socio-economic Status
SLI: Standard of Living Index
TV: Television

## Declarations

### Ethics approval and consent to participate

Ethics approval for the study was obtained from the Ethics Committee of KEM Hospital Research Centre and all participants gave informed consent for this study.

### Consent for publication

Not applicable

## Competing interests

The authors declared that they had no competing interests.

## Funding

This study was supported by The Wellcome Trust, London, U.K., the U.K. Medical Research Council, and the Department of Biotechnology, Government of India.

## Authors’ contributions

CSY and CHDF jointly conceptualised and started the study in 1993, and its ongoing data collection. They also guided the analysis, and edited the manuscript. S.R.S. performed statistical analysis and wrote the manuscript. K.K. interacted with SRS for analysis, wrote, and edited the manuscript. AR helped to draft the manuscript. S.D.C, T.M.D, A.J.B and V.A.S collected the data. PCY monitored the study data collection.

C.S.Y. is the guarantor of this work and, as such, has full access to all the data in the study and takes responsibility for the integrity of the data and the accuracy of the data analysis.

## Acknowledgements

The authors are grateful to the study participants for taking part in this study. The authors thank the late Dr. B.J. Coyaji and Dr. V.N. Rao, KEM Hospital Research Center, Pune, India, and Dr K J Coyaji for providing research facilities. The authors also acknowledge the support of SNEHA-India.

## Notes

### Competing Interest Statement

The authors have declared no competing interest.

