## Additional tables for "Socio-economic Transition in an Indian Rural Setting and Its Impact on Health: Pune Maternal Nutrition Study"

**Supplementary Table 1: Primary census abstract of study villages along with amenities available, 1991-2001.**

| **Village** | **Dhamari** | | **Pabal** | | **Karandi** | | **Kendur** | | **Shikrapur** | | **Pimple Jagtap** | |
| --- | --- | --- | --- | --- | --- | --- | --- | --- | --- | --- | --- | --- |
| Year | 1991 | 2001 | 1991 | 2001 | 1991 | 2001 | 1991 | 2001 | 1991 | 2001 | 1991 | 2001 |
| Code | 36 |  | 22 |  | 41 |  | 32 |  | 70 |  | 68 |  |
| **Amenities available** |  |  |  |  |  |  |  |  |  |  |  |  |
| Educational | P,M,H | P,M,H | P,M | P,M,H,PUC | P,M,H | P,M,H | P,M,H,PUC | P,M,H,PUC | P,M,H,PUC | P,M,H,PUC | P,M | P,M,H |
| Medical | - (-5) | MCW,PHC(5-10) | HC,PHC,H,PUC | MCW | -(5-10) | -(10+) | MCW,CWC,PHC,D,FPC | MCW,CWC,PHC,D,FPC | D(2)RP | MCW (10+) | HC | HC(5-10), MCW (5-10), PHC (5-10) |
| Drinking water potable | T,W | T,W,HP | T,W,R | T,W,HP | T,W | T.W.HP,TW | T,W,R | T,W,HP,R | W,HP,R | T,W,TW,HP, R | T,W | T, W,TW, HP |
| Post and telegraph | PO, Phone | PTO, Phone | PTO, Phone | PTO, Phone | PO | PTO, Phone | PO, Phone | PTO, Phone | PO, Phone | PO, Phone | -(-5) | PO (10+), Phone |
| Day or days of the market | - (5-10) |  | Fri |  | -(10+) |  | Mon |  | -(-5) |  | -(10+) |  |
| Communication (Bus stop, railway station, water way) | BS | BS,RS (10+) | BS | BS, RS (10+) | BS | BS, RS(10+) | BS | BS, RS (10+) | BS | BS, RS (10+) | BS | BS, RS (10+) |
| Approach to village | PR | PR | PR | PR | PR | PR, Canal | PR | PR | PR | PR | PR | PR |
| Power supply | EA | EA | EA | EA | EA | EA | EA | EA | EA | EA | EA | EA |
| **Land used (area under different, types of land use in hectares rounded to two decimal places)** | | | | | | | |  |  |  |  |  |
| Forest | 149.00 | 149.00 | 521.00 | 501.00 | 285.00 | 127.00 | - | 00 | - | 0.00 | - | 0.00 |
| Irrigated by source | WE (270.00) | WE (529.00) | WE (465.00) | WE (650.00) | WE (170.00) | WE (873.00) | WE (326.00) | WE (668.00) | WE (260.00) | WE (75.00) | WE (120.00) | WE (121.00) |
| Unirrigated | 2763.00 | 2664.00 | 2406.00 | 2253.00 | 1658.00 | 1093 | 3058.00 | 3078.00 | 1754.00 | 1710.53 | 1455.00 | 611.58 |
| Culturablewaste (including gauchar and groves) | 117.00 | 80.00 | 96.00 | 275.00 | 300.00 | 52.00 | 307.00 | 306 | 96.00 | 305.00 | 8.00 | 130.00 |
| Area not available for cultivation | 268.00 | 145.00 | 490.00 | 299.00 | 129.00 | 398.00 | 705.00 | 344.00 | 139.00 | 158.41 | 286.00 | 235.38 |
| **Other facility** |  |  |  |  |  |  |  |  |  |  |  |  |
| Banking facility | NA | Yes |  | Yes |  | Yes |  | Yes |  | Yes |  | NA(10+) |
| Credit societies | NA | Yes |  | Yes |  | Yes |  | Yes |  | Yes |  | Yes |
| Recreational and cultural facilities | NA | Yes |  | NA |  | NA |  | NA |  | NA (10+) |  | NA (10+) |
| Newspaper/magazine | NA | Yes |  | Yes |  | Yes |  | Yes |  | Yes |  | Yes |
| **Total income and expenditure of village (in Rs’00)** | NA | Yes |  | Yes |  | Yes |  | Yes |  | Yes |  | Yes |
| Total income |  | 304 |  | 522 |  | 4435 |  | 442 |  | 12329 |  | 1105 |
| Total expenditure |  | 244 |  | 393 |  | 3750 |  | 441 |  | 10809 |  | 1108 |

- P=Primary, M=Junior secondary or middle, H=Matriculate or secondary school, PUC=Higher secondary or pri-university or junior college
- H=Hospital, MCW=Maternity and child welfare center, CWC=Child welfare center, PHC=Primary health cedenter, D=Dispensary, FPC=Family planning center, RP=Registered private practitioner, HC=Health centre
- T=Tap water, W= Well water, R=River water, HP=Hand pump, TW=Tube well
- PO=Post office, TO=Telegraph office, PTO=Post and telegraph office, Phone= Telephone connection
- BS=Bus stop, RS=Railway station
- PR=Pucca road (cement concrete, Black top water bound Macadam)
- EA=Electricity for domestic, agriculture, industrial or commercial purpose
- WE=Well (with electricity)

**Supplementary Table 2**: **Child anthropometry, biochemistry and nutritional intake at 12y follow-up by change (6y to 12y)in Standard of Living Index according to gender.**

|  | **Change in SLI from 6 to 12y- BOYS-N=285** | | | | | | **Change in SLI from 6 to 12y- GIRLS-N=285** | | | | | |
| --- | --- | --- | --- | --- | --- | --- | --- | --- | --- | --- | --- | --- |
| **At 12y anthropometry** | **No transition**  **(90)** | **Below average transition (119)** | **Above average transition**  **(76)** | **p** | **P1 (Age, birth-order)** | **P2 (Age, birth-order, Migration)** | **No transition**  **(100)** | **Below average transition (93)** | **Above average transition**  **(92)** | **p** | **P1 (Age, birth-order)** | **P2 (Age, birth-order, Migration)** |
| Age (y) | 11.7 (0.9) | 11.5 (0.9) | 11.9 (0.9) | 0.387 | -- | -- | 11.5 (0.9) | 11.5 (0.9) | 11.8 (0.8) | 0.052 | -- | -- |
| Weight (kg) | 29.5 (5.9) | 27.8 (4.9) | 31.1 (7.7) | 0.141 | 0.031 | 0.234 | 29.5 (6.9) | 29.0 (7.4) | 30.7 (8.4) | 0.271 | 0.950 | 0.951 |
| Weight gain from birth till 12y | 27.0 (5.8) | 25.1 (4.6) | 28.8 (7.8) | 0.110 | 0.018 | **0.010** | 26.7 (6.8) | 26.3 (7.1) | 28.4 (8.4) | 0.135 | 0.922 | 0.918 |
| Height (cm) | 140.2 (8.9) | 136.8 (7.5) | 141.6 (9.1) | 0.409 | 0.009 | **0.013** | 139.6 (8.2) | 139.9 (8.7) | 141.6 (8.3) | 0.106 | 0.815 | 0.818 |
| Body mass index (kg/m2) | 14.9 (1.6) | 14.7 (1.4) | 15.3 (2.2) | 0.143 | 0.310 | 0.312 | 14.9 (2.1) | 14.6 (2.4) | 15.1 (2.6) | 0.684 | 0.677 | 0.696 |
| Waist-hip ratio | 0.85 (0.04) | 0.88 (0.37) | 0.86 (0.37) | 0.842 | 0.494 | 0.484 | 0.81 (0.06) | 0.95 (1.4) | 0.80 (0.05) | 0.974 | 0.269 | 0.272 |
| Head circumference (cm) | 51.5 (1.7) | 51.4 (1.4) | 51.6 (1.7) | 0.796 | 0.998 | 0.998 | 50.9 (1.8) | 51.2 (2.1) | 51.2 (1.9) | 0.405 | 0.555 | 0.558 |
| SS/TR ratio | 0.86 (0.18) | 0.82 (0.15) | 0.86 (0.16) | 0.692 | 0.170 | 0.250 | 0.95 (0.20) | 0.92 (0.19) | 0.97 (0.22) | 0.514 | 0.423 | 0.419 |
| Total fat percent | 14.7 (6.7) | 14.7 (4.2) | 15.9 (7.7) | 0.224 | 0.439 | 0.434 | 19.3 (6.2) | 18.6 (6.5) | 19.6 (7.6) | 0.764 | 0.713 | 0.741 |
| Total lean percent | 80.9 (6.5) | 80.9 (4.2) | 79.7 (7.5) | 0.227 | 0.433 | 0.428 | 76.4 (6.1) | 77.1 (6.4) | 76.1 (7.5) | 0.800 | 0.732 | 0.760 |
| **Blood pressure (mmHg)** |  |  |  |  |  |  |  |  |  |  |  |  |
| Systolic/Diastolic | 106/62 (9/7) | 106/63 (11/7) | 107/63 (11/6) | 0.331,0.401 | 0.666, 0.735 | 0.537, 0.571 | 105/62 (11/7) | 104/62 (10/7) | 106/63 (11/7) | 0.303, 0.126 | 0.885, 0.414 | 0.888, 0.433 |
| **Hemogram** |  |  |  |  |  |  |  |  |  |  |  |  |
| Hemoglobin (g/dL) | 12.7 (0.9) | 13.0 (1.0) | 12.9 (1.0) | 0.270 | 0.285 | 0.190 | 12.6 (1.1) | 12.7 (1.1) | 12.8 (0.9) | 0.294 | 0.367 | 0.356 |
| WBC count | 7.1 (1.8) | 7.3 (2.0) | 7.7 (2.3) | 0.099 | 0.084 | **0.027** | 7.2 (1.7) | 7.8 (2.1) | 7.2 (1.6) | 0.826 | 0.092 | 0.092 |
| **Glucose (mg%)** |  |  |  |  |  |  |  |  |  |  |  |  |
| Fasting | 87.8 (6.7) | 87.3 (6.9) | 88.3 (6.9) | 0.689 | 0.344 | 0.555 | 85.6 (8.2) | 85.7 (5.9) | 86.5 (7.8) | 0.436 | 0.554 | 0.539 |
| **Insulin (mU/L)** |  |  |  |  |  |  |  |  |  |  |  |  |
| Fasting | 5.1 (2.9) | 5.2 (3.1) | 5.2 (2.7) | 0.861 | 0.916 | 0.922 | 6.5 (4.1) | 5.8 (2.9) | 7.0 (3.6) | 0.391 | 0.193 | 0.199 |
| **Lipids** |  |  |  |  |  |  |  |  |  |  |  |  |
| Triglycerides | 56.8 (18.8) | 55.0 (19.0) | 62.0 (21.2) | 0.112 | 0.119 | 0.321 | 60.2 (24.1) | 58.9 (23.5) | 65.3 (27.7) | 0.172 | 0.295 | 0.270 |
| Total cholesterol | 127.8 (23.1) | 131.9 (21.4) | 135.9 (26.0) | 0.027 | 0.101 | 0.316 | 132.0 (23.5) | 129.3 (19.0) | 135.3 (21.5) | 0.303 | 0.247 | 0.243 |
| HDL cholesterol | 41.3 (8.7) | 42.2 (8.6) | 41.8 (8.3) | 0.719 | 0.913 | 0.882 | 42.3 (9.6) | 42.0 (7.5) | 42.9 (9.5) | 0.656 | 0.579 | 0.583 |
| **Total intake** |  |  |  |  |  |  |  |  |  |  |  |  |
| Caloric | 2246.3 (741.3) | 2066.8 (636.7) | 2116.7 (611.9) | 0.190 | 0.271 | 0.165 | 1864.9 (548.6) | 1870.4 (634.1) | 1863.7 (473.8) | 0.989 | 0.946 | 0.894 |
| Carbohydrate | 397.3 (132.6) | 369.8 (116.6) | 375.3 (105.8) | 0.218 | 0.349 | 0.188 | 329.2 (92.5) | 328.3 (106.4) | 327.9 (82.5) | 0.929 | 0.955 | 0.918 |
| Fat | 44.6 (18.0) | 10.0 (15.2) | 42.7 (17.4) | 0.430 | 0.324 | 0.379 | 37.6 (15.4) | 38.5 (17.5) | 37.7 (12.7) | 0.930 | 0.811 | 0.769 |
| Protein | 60.6 (21.0) | 55.7 (17.7) | 56.9 (17.4) | 0.192 | 0.292 | 0.151 | 49.9 (15.1) | 50.3 (17.9) | 50.9 (14.0) | 0.691 | 0.979 | 0.920 |

Values are mean (SD; P is significance value for trend by ANOVA; P1 adjusted for age of the child at the time of measurements; P2 adjusted for age of the child, birth order.

**Supplementary Table 3: Child anthropometry, biochemistry and nutritional intake at 18y follow-up by change (12y to 18y) inStandard of Living Index according to gender.**

|  | **Change in SLI from 12 to 18y- BOYS-N=285** | | | | | | **Change in SLI from 12 to 18y- GIRLS-N=272** | | | | | |
| --- | --- | --- | --- | --- | --- | --- | --- | --- | --- | --- | --- | --- |
| **At 18y anthropometry** | **No transition**  **(52)** | **Below average transition (84)** | **Above average transition**  **(149)** | **p** | **P1 (Age)** | **P2 (Age, birth-order)** | **No transition**  **(74)** | **Below average transition (79)** | **Above average transition**  **(117)** | **p** | **P1 (Age)** | **P2 (Age, birth-order)** |
| Age (y) | 18.2 (0.5) | 18.2 (0.5) | 18.2 (0.5) | 0.847 | -- | -- | 17.6 (0.6) | 17.7 (0.6) | 17.7 (0.5) | 0.632 | -- | -- |
| Weight (kg) | 56.3 (10.2) | 56.4 (12.3) | 55.9 (10.2) | 0.796 | 0.958 | 0.957 | 45.9 (7.9) | 47.1 (7.7) | 46.6 (7.9) | 0.609 | 0.672 | 0.662 |
| Weight gain from birth till 18y | 53.7 (10.2) | 53.5 (12.3) | 53.4 (10.3) | 0.906 | 0.970 | **0.968** | 43.5 (7.9) | 44.6 (7.6) | 43.8 (7.9) | 0.893 | 0.728 | 0.728 |
| Height (cm) | 170.5 (6.1) | 169.2 (7.3) | 168.8 (6.7) | 0.124 | 0.239 | **0.239** | 157.2 (6.6) | 156.9 (6.2) | 156.9 (5.2) | 0.789 | 0.943 | 0.969 |
| Body mass index (kg/m2) | 19.3 (3.2) | 19.6 (3.8) | 19.6 (2.9) | 0.701 | 0.906 | 0.906 | 18.5 (3.0) | 19.1 (3.1) | 18.9 (3.3) | 0.499 | 0.555 | 0.571 |
| WHR | 0.82 (0.06) | 0.84 (0.16) | 0.83 (0.05) | 0.755 | 0.328 | 0.320 | 0.78 (0.04) | 0.79 (0.05) | 0.79 (0.04) | 0.663 | 0.408 | 0.373 |
| Head circumference (cm) | 53.8 (1.4) | 53.6 (1.9) | 53.9 (1.8) | 0.493 | 0.362 | 0.363 | 52.6 (1.4) | 52.5 (1.4) | 52.7 (1.2) | 0.380 | 0.427 | 0.429 |
| Total fat percent | 14.2 (8.2) | 16.1 (8.5) | 15.5 (8.2) | 0.471 | 0.428 | 0.432 | 27.7 (7.4) | 28.6 (6.3) | 28.3 (6.9) | 0.590 | 0.748 | 0.804 |
| Total lean percent | 81.5 (8.0) | 79.6 (8.3) | 80.3 (7.9) | 0.520 | 0.431 | 0.435 | 68.0 (7.3) | 67.1 (6.1) | 67.4 (6.6) | 0.541 | 0.735 | 0.791 |
| **Blood pressure (mmHg)** |  |  |  |  |  |  |  |  |  |  |  |  |
| Systolic/Diastolic | 111/57 (12/8) | 113/60 (11/9) | 112/59 (10/8) | 0.639, 0.571 | 0.500, 0.182 | 0.501, 0.183 | 106/62 (7/6) | 104/61 (8/6) | 105/61 (9/7) | 0.924, 0.356 | 0.582, 0.517 | 0.567, 0.460 |
| **Hemogram** |  |  |  |  |  |  |  |  |  |  |  |  |
| Heamoglobin (g/dL) | 13.9 (1.2) | 14.0 (1.3) | 14.0 (1.1) | 0.451 | 0.714 | 0.715 | 11.8 (0.9) | 11.6 (1.2) | 11.7 (1.2) | 0.682 | 0.590 | 0.608 |
| WBC count | 5.9 (1.3) | 6.1 (1.3) | 5.8 (1.4) | 0.426 | 0.231 | **0.232** | 6.3 (1.3) | 6.7 (1.6) | 6.6 (1.5) | 0.371 | 0.227 | 0.235 |
| **Glucose (mg%)** |  |  |  |  |  |  |  |  |  |  |  |  |
| Fasting | 97.6 (5.9) | 95.7 (6.0) | 97.6 (6.4) | 0.493 | 0.542 | 0.541 | 93.5 (6.2) | 92.4 (5.4) | 92.6 (5.8) | 0.320 | 0.320 | 0.377 |
| 2h | 111.5 (20.1) | 112.6 (26.8) | 109.9 (20.1) | 0.518 | 0.859 | 0.860 | 111.9 (20.7) | 114.4 (22.0) | 113.6 (22.6) | 0.692 | 0.755 | 0.822 |
| **Insulin (mU/L)** |  |  |  |  |  |  |  |  |  |  |  |  |
| Fasting | 9.4 (5.2) | 10.6 (5.4) | 10.4 (6.8) | 0.451 | 0.542 | 0.541 | 11.2 (4.1) | 11.0 (4.4) | 11.2 (4.7) | 0.908 | 0.972 | 0.973 |
| **Lipids** |  |  |  |  |  |  |  |  |  |  |  |  |
| Triglycerides | 65.6 (21.3) | 65.6 (27.6) | 65.8 (31.1) | 0.955 | 0.998 | 0.998 | 55.7 (17.2) | 57.6 (19.9) | 59.1 (23.5) | 0.281 | 0.551 | 0.564 |
| Total cholesterol | 124.7 (24.1) | 127.3 (25.2) | 123.7 (22.1) | 0.555 | 0.511 | 0.512 | 133.4 (21.1) | 129.6 (21.2) | 133.8 (23.8) | 0.769 | 0.420 | 0.435 |
| HDL cholesterol | 37.3 (7.6) | 39.8 (11.4) | 38.7 (11.9) | 0.664 | 0.410 | 0.410 | 44.5 (8.6) | 41.6 (8.0) | 42.3 (9.9) | 0.147 | 0.107 | 0.124 |
| HOMA-beta | 99.2 (32.3) | 111.5 (37.4) | 107.4 (48.4) | 0.417 | 0.177 | 0.178 | 122.9 (31.7) | 125.0 (32.9) | 125.2 (35.0) | 0.683 | 0.917 | 0.909 |
| HOMA-IR | 1.41 (0.75) | 1.56 (0.77) | 1.62 (1.26) | 0.249 | 0.757 | 0.758 | 1.6 4(0.59) | 1.63 (0.62) | 1.65 (0.67) | 0.935 | 0.917 | 0.909 |
| **Total intake (data not calculated)** |  |  |  |  |  |  |  |  |  |  |  |  |

Values are mean (SD; P is significance value for trend by ANOVA; P1 adjusted for age of the child at the time of measurements; P2 adjusted for age of the child, birth order.

**Supplementary Table 4: Maternal anthropometry, biochemistry and nutritional intake at 12y follow-up by change (6-12y) in Standard of Living Index according to gender.**

|  | **Change in SLI from 6 to 12y- mother (N=570)** | | | | | |
| --- | --- | --- | --- | --- | --- | --- |
| **At 12y anthropometry** | **No transition**  **(186)** | **Below average transition (208)** | **Above average transition(158)** | **p** | **p1** | **p2** |
| Age (y) | 33.3 (3.3) | 33.0 (3.5) | 33.5 (3.5) | 0.492 | -- |  |
| Weight (kg) | 47.7 (8.5) | 45.9 (7.8) | 49.1 (8.7) | 0.174 | **0.004** | **0.0001** |
| Height (cm) | 153.4 (5.4) | 152.5 (4.8) | 153.6 (5.1) | 0.756 | 0.072 | **0.051** |
| Body mass index (kg/m2) | 20.3 (3.5) | 19.7 (3.1) | 20.7 (3.4) | 0.204 | **0.024** | **0.003** |
| **Blood pressure (mmHg)** |  |  |  |  |  |  |
| Systolic/ Diastolic | 104/62 (10/9) | 102/60 (8/7) | 103/62 (10/9) | 0.351, 0.510 | 0.065, 0.097 | 0.115, 0.107 |
| **Hemogram** |  |  |  |  |  |  |
| Heamoglobin (g/dL) | 11.8 (1.8) | 11.9 (1.9) | 12.2 (1.5) | **0.039** | 0.097 | 0.082 |
| WBC count | 6.9 (1.9) | 6.7 (1.6) | 6.9 (1.5) | 0.844 | 0.461 | 0.435 |
| Creatinine (mg%) | 0.79 (0.13) | 0.79 (0.14) | 0.83 (0.14) | 0.083 | **0.021** | **0.028** |
| **Glucose (mg%)** |  |  |  |  |  |  |
| Fasting | 89.4 (12.9) | 89.8 (14.6) | 90.1 (9.4) | 0.632 | 0.931 | 0.964 |
| **Insulin (mU/L)** |  |  |  |  |  |  |
| Fasting | 5.9 (3.9) | 5.6 (2.9) | 6.1 (3.8) | 0.688 | 0.466 | 0.191 |
| HOMA-R | 1.30 (0.90) | 1.27 (0.74) | 1.39 (0.96) | 0.408 | 0.638 | 0.189 |
| **Lipids** |  |  |  |  |  |  |
| Triglycerides | 69.3 (35.1) | 67.2 (32.4) | 71.6 (29.2) | 0.552 | 0.203 | **0.057** |
| Total cholesterol | 151.1 (30.9) | 147.5 (30.8) | 156.3 (30.3) | 0.148 | 0.020 | **0.003** |
| HDL cholesterol | 44.4 (9.5) | 44.2 (9.4) | 43.9 (7.9) | 0.618 | 0.893 | 0.973 |
| **Folate-B12** |  |  |  |  |  |  |
| Serum folate (nmol/L) | 20.7 (13.3) | 20.5 (10.2) | 21.5 (13.0) | 0.574 | 0.718 | 0.813 |
| Vitamin B_12_ (pmol/L) | 216.3 (253.7) | 203.5 (174.5) | 180.7 (74.2) | 0.082 | 0.189 | 0.335 |
| Homocystein (µM/L) | 15.4 (8.3) | 15.5 (9.9) | 16.6 (6.4) | 0.416 | 0.184 | 0.080 |

Values are mean (SD; P is significance value for trend by ANOVA; P1 adjusted for age of the child at the time of measurements; P2 adjusted for age of the child, birth order.
