## Additional Fig for "Socio-economic Transition in an Indian Rural Setting and Its Impact on Health: Pune Maternal Nutrition Study"

**Supplementary Fig 1: Development parameters in the study area at the 6 (2000-2), 12 (2006-8) and 18 years (2013-14) follow-up compared with rural Maharashtra at the NFHS II (1998-9) and NFHS III (2005-6) surveys**

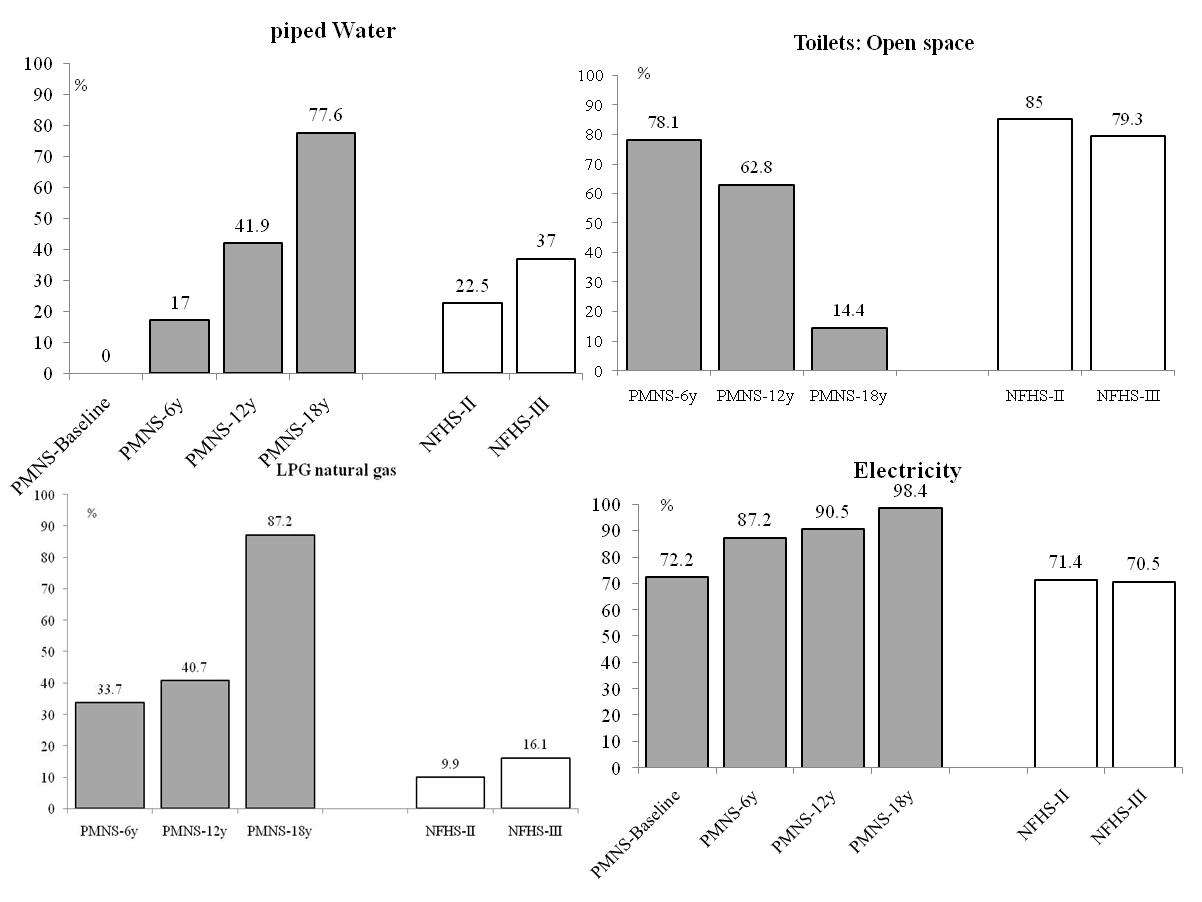

Collected SLI data used for study area; NFHS data used for rural Maharashtra.
